# Reliability and Predictive Validity of a Gait Assessment using Inertial Measurement Units: The Importance of Standardizing Walking Surface and Footwear

**DOI:** 10.1101/2022.03.17.22272451

**Authors:** Len Lecci, Kelly Dugan, Ken Zeiger, Julian Keith, Sasidharan Taravath, Wayland Tseh, Mark Williams

## Abstract

**Objectives:** Evaluate procedures for analyzing raw accelerometer data (inertial measurement units) to reconstruct the gait cycle using BioKinetoGraph (BKG). We examine whether footwear and walking surface influence gait (BKG) and evaluate test-retest reliability. We also examine the association between BKG and NIH 4-meter gait, and compare BKG to other neurobehavioral measures for predicting concussion symptoms.

**Methods:** In Study 1, a within-subjects design with 60 participants was used to examine the effects of footwear (shoes/no-shoes) and walking surface (tile floor/grass) on BKG data, and evaluate retest reliability. Study 2 employed a cross-sectional, cohort design of 1,008 participants to assess BKG’s correlation with NIH 4-m gait, and prediction of Centers of Disease Control and Prevention (CDC) concussion symptoms relative to previously validated speed and balance measures.

**Results:** 2×2 ANOVAs illustrate footwear and walking surface effects on BKG for the power, stride, stability, and symmetry, with variable effect sizes. Retest reliability (Pearson *r*s) for the no shoes/ tile surface condition ranged from .72-.91 (mean = .80, 4-day average interval). BKG correlates significantly with NIH 4-m gait. Regression analyses found BKG predicts CDC concussion symptom endorsement, and outperforms (2-3 fold) BESS and NIH 4-meter gait.

**Conclusions:** Gait assessments should be standardized for footwear and especially walking surface. When standardized (no shoes/hard surface) BKG results in strong test-retest reliability. BKG variables are strongly related to NIH 4-m gait, and are superior to standard measures of gait speed and balance when predicting concussion symptoms; offering additional information when predicting the sequalae of concussion.

**Summary Box:** *What is already known on the topic?:*

- *Sensor technology to evaluate gait has established reliability and predicts a wide range of medical outcomes. However, the influence of footwear and walking surface on gait has not been studied, nor has a sensor-based gait assessment been compared to conventional measures for predicting concussion symptoms*.

*What this study adds?:*

- *Gait sensor data is sensitive to footwear and walking surface, but can produce good reliability when these factors are standardized. Gait sensor scores converge with other validated measures of gait, and sensor-based measures of power, stride, symmetry, and stability can outperform established gait speed and balance measures when predicting CDC concussion symptoms*.

*How this study might affect research, practice, or policy?:*

- *Uniform standards for footwear and walking surface are needed when evaluating gait in both research and practice, and sensor-based gait measures can be reliably assessed to provide insight into the behavioral sequelae of concussion, that are superior to simple gait speed*.

## INTRODUCTION

The interpretation of human movement plays an essential role in the clinical practice of numerous medical specialties, including pediatrics, sports medicine, geriatrics, neurology, rheumatology, orthopedics, and rehabilitation. Even walking speed can predict important outcomes, including response to rehabilitation, functional dependence, fatality, mobility, cognitive decline, and hospitalizations [1]. To date, most work has focused on geriatric populations [2], and recommendations from the *Mobility Working Group* emphasize gait speed as chief predictor of mobility and various life outcomes [3].

Advanced instrumentation to capture and record elements of gait functioning can add significantly to the understanding of gait [4]. For example, gait speed assessed using accelerometers has superior sensitivity to dysfunction relative to manually collected data [5]. Accelerometers can produce reliable outputs across a range of clinical populations [6-12]. However, the specific influence of footwear and walking surface on accelerometers has received little attention. Variability in footwear and walking surface may influence movement biomechanics [13], and gait outcomes in adults [14] and children [15], and if unstandardized, these factors could introduce measurement error, thereby lowering reliability and validity.

The current research uses noninvasive, continuous triaxial accelerometer data to create a BioKinetoGraph (BKG). Analogous to the 12-lead electrocardiogram, BKG waveforms are related to specific neuromotor gait cycle events, including cadence, foot-strike, push-off, double stance time, and swing phase. Raw data are combined to generate BKG waveforms as gravitational accelerations over time. Tracings are used to obtain range of motion, amplitudes, and timing intervals for the various components of the waveform that relate directly to the gait cycle. Figure 1 illustrates the BKG from one ankle accelerometer and its associated components of the gait cycle. Simultaneously recorded high-speed videos validated the BKG waves to the corresponding gait action.

**Figure 1.**
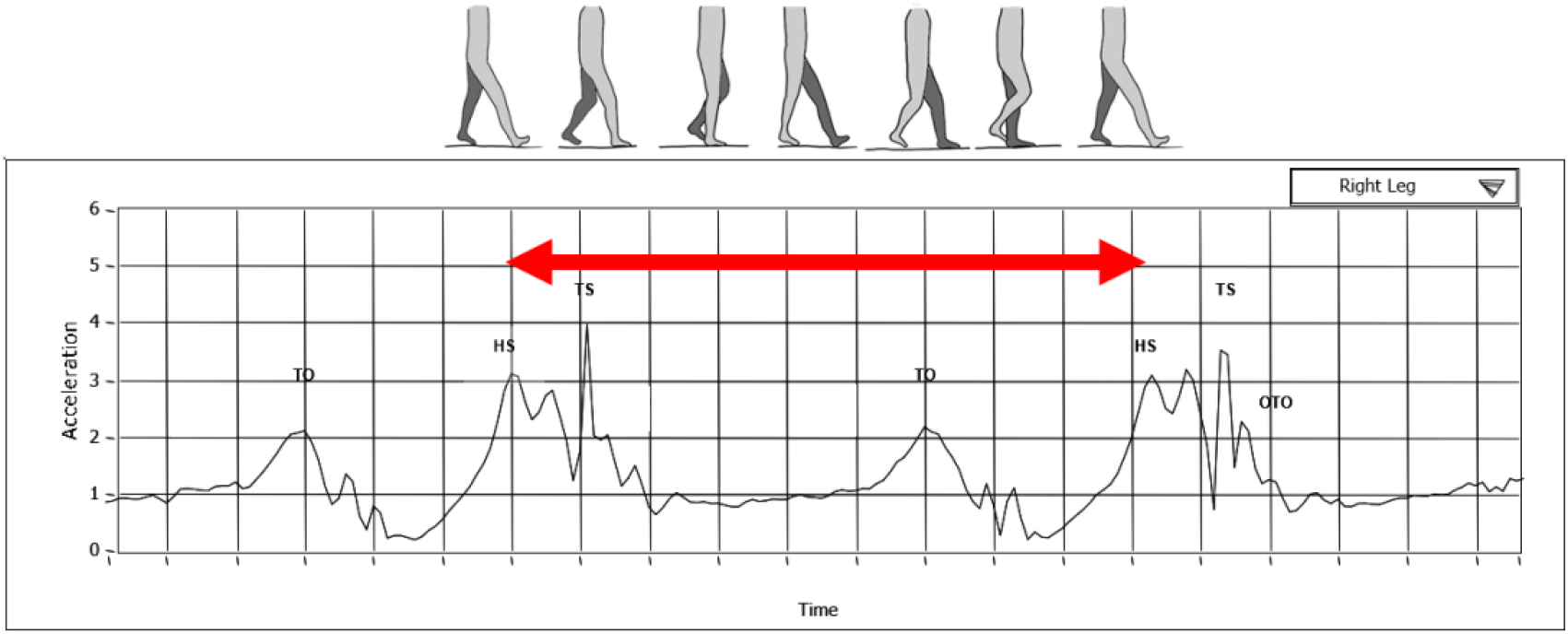
Illustration of a 3 second BKG signature drawn from a single (right ankle). The arrow shows one heel strike interval for one step. TO represents toe off. HS represents heel strike. TS represents toe strike. Clinical conditions can alter the size, shape, and timing of the waves.

Two studies are presented with objectives of examining the effects of walking surface and footwear on BKG gait, establishing BKG test retest reliability, convergent validity with the NIH 4-meter gait, and comparative validity of the BKG relative to conventional gait speed and balance measures when predicting self-reported CDC concussion symptoms.

## STUDY 1

Aims to examine the influence of two extraneous variables (footwear and walking surface) on BKG, and evaluate BKG test-retest reliability.

### Methods

#### Patient and Public Involvement

Volunteers agreed to have their gait evaluated while systematically varying footwear and walking surface. Participants were informed of the goal to evaluate sensor capabilities and educated on the benefits of research, but were not otherwise involved. UNCW’s IRB approved studies 1 and 2. No adverse outcomes were reported.

#### Data Availability Statement

Deidentified data and protocol (for studies 1 and 2) may be obtained by providing methodologically sound proposals. Direct inquires to.

#### Procedure

Employed a within-subjects design, with order counterbalanced. Those enrolled in physical education classes at a university in the southeastern United States and ambulating independently, were eligible. Researchers and participants were aware of the footwear and walking surface, but blind to BKG outcomes.

The gait protocol involves starting at one end of a 20-foot straight walking path, walking at their usual pace, turning, and walking back to the starting point.

##### Day 1

Participants wearing a sacrum gait sensor performed the above gait protocol covering 20 feet and back on an even, hard surface (tile floor), then on an uneven surface (grass), with and without shoes on each surface. Participants then repeated each condition (eight trials total, counterbalancing order).

##### Day 2

Repeated the Day 1 protocol, with retest intervals ranging from 1 to 14 days (Mean = 4.04, SD = 2.68), with 95% completed within 9 days. Presented values reflect the mean of two walking trials under the same conditions on the same day. If a second trial was unavailable, which occurred for 10% of participants, single trial data were used. Other missing data resulted in case-wise exclusion.

Although the full BKG is evaluated from sensors located at each ankle, sacrum, and wrist, we report the single sacrum sensor (near the small of the back). Sensors record inertial measurement unit (IMU; a general term for accelerometers, gyroscopes, and similar recording devices), via transmitted signals at a rate of 200 Hz to a local Microsoft Surface Pro, which transmits the data to remote servers for BKG motion analysis following each trial. Raw data are saved to a persistent data store as a collection of timestamps and accelerometer readings of each axis. Raw inertial data are then processed through a series of algorithms to detect key markers of the gait cycle and extract the spatial, temporal, kinetic, and spectral features reported herein.

Prior to detecting the initial contact (heel-strike) and final contact (toe-off) events of each gait cycle, the signal is processed to remove extraneous data recorded before and after the gait trial, adjust for tilt and orientation of the sensor relative to gravity, and filtered to reduce noise. Correct orientation is ensured using template correlation with the y-axis pointed downward, the x-axis pointed right, and the z-axis rearward. Tilt due to the placement of the sensor on the body is adjusted along each axis so that the vertical axis is centered at zero. The raw signal data is then fourth-order-zero-phase-shift filtered [16] with an upper cutoff of 3hz and a lower cutoff of 0.111hz. Initial contact and final contact events were detected algorithmically by first identifying local maxima representative of the mid-swing with toe-off and heel strike events indicated by minima occurring immediately before and after, respectively. Displacement and velocity were obtained for each axis by trapezoidal integration and subtraction of the zero-phase rolling average [17] equal to one gait cycle.

BKG output represents sixteen variables, organized into four conceptual domains of stability, timing intervals (stride), amplitudes (power), and regularity (symmetry).

Stability is range of motion (ROM) at the center of mass (sacrum) as well as gait cycle variability, represented by variables that occur during the straight portion of the gait cycle (sway) and those occurring while turning around (turnaround sway), the velocity of side to side movement (sway velocity), the rhythmicity of gait patterns (gait smoothness), and the variability of time in double stance phase (support variability).

Stride refers to maintaining velocity during walking and includes the average timing interval between contralateral heel strikes (stride time) and the average duration spent in the double stance phase (double stance time).

Symmetry refers to consistency of movement across the anterior-posterior and vertical dimensions. This includes a comparison of the forward and backward displacement from the center of mass (forward movement symmetry), a comparison of the distance of upward and downward displacement from the center of mass (vertical movement symmetry), and a comparison of the velocity of upward and downward movement from the center of mass (vertical sway symmetry).

Power refers to amount and efficiency of energy generated and expended in the gait cycle. This includes total net force generated at the COM (total power), force generated with vertical movement (vertical power), force produced with forward movement (forward power), force generated with lateral movement (side power), force generated by the foot strike (striking force), and force generated by the toe pushing off (pushing force).

Effects of surface type (grass or tile floor), and footwear (shoes or no shoes) on the raw scores of each of the sixteen BKG variables were assessed using 2 × 2 ANOVAs. Retest reliability was assessed using Pearson correlations between BKG values from Days 1 and 2.

A target sample size was set at > 48 to detect a small to medium effect size (d = .35) with power = .80.

### Results

#### Participants

In the Fall of 2016, college students with no obvious health problems, aged 18 to 35 (Mean = 21.26 ± 2.79 years), 61.7% female (Height = 64.79 ± 12.87 inches; Weight = 149.80 ± 37.11 lbs.), were recruited and consented prior to initiating the procedure. All participant data are included, aside from those whose sensors malfunctioned. Evaluating the effects of walking surface and footwear on gait accelerometer data

Significant and relatively large main effects were observed for surface type on the power domain variables of total power, vertical power, side power, forward power, pushing force, and striking force, as well as stride time, forward movement symmetry, gait smoothness, sway velocity and turnaround sway (Table 1, Supplemental material includes significant ANOVAs). These main effects indicate that when walking on grass, compared to tile, participants expended more energy across all axes, had longer stride times, exhibited less symmetry in forward and backward movement, displayed irregular gait patterns, showed greater side to side sway velocity, and manifested greater sway during the turnaround.

**Table 1.** Pearson Correlations Illustrating Test-Retest Reliabilities for BKG Variables by Domain

| Domain | Variable | Reliability<br>(*r*) | Correlation with<br>NIH 4M Gait |
| --- | --- | --- | --- |
| Power | *Reliability $r_{average}$*<br>*= .79* |  |  |
|  | 1. Total power | .71** | -.18** |
|  | 2. Forward power | .75** | -.29** |
|  | 3. Side power | .83** | -.35** |
|  | 4. Vertical power | .80** | -.45** |
|  | 5. Pushing force | .79** | -.30** |
|  | 6. Striking force | .83** | -.30** |
| Stride | *Reliability $r_{average}$*<br>*= .86* |  |  |
|  | 7. Stride time | .91** | .31** |
|  | 8. Double stance | .81** | .25** |
| Stability | *Reliability $r_{average}$*<br>*= .81* |  |  |
|  | 9. Support variability | .82** | .17** |
|  | 10. Gait smoothness | .72** | -.06* |
|  | 11. Sway | .89** | .00 |
|  | 12. Turnaround sway | .76** | .06 |
|  | 13. Sway velocity | .88** | -.11** |
| Symmetry | *Reliability $r_{average}$*<br>*= .75* |  |  |
|  | 14. Forward<br>Movement Symmetry | .76** | .09** |
|  | 15. Vertical<br>Movement Symmetry | .78** | -.05 |
|  | 16. Vertical Sway<br>Symmetry | .67** | .33** |

*Note.* \* $p < .05$ , \*\* $p < .01$ . Cumulatively, BKG variables explain 25.6% variance (multiple $R = .51$ ) of NIH 4 m gait scores ( $F = 21.4$ , $df = 16, 993$ , $p < .001$ ).

Significant, but small to medium main effects of footwear were observed for vertical power, striking force, stride time, double stance time, and vertical sway symmetry. This indicates that when wearing shoes, compared to no shoes, participants displayed longer stride times, longer double stance times, greater symmetry in the speed of upward and downward movement, greater energy expenditure with vertical movement, and greater energy expenditure when the foot strikes the walking surface. No interaction effects emerged.

#### Reliability of accelerometer gait assessment

BKG test-retest reliability was examined by holding constant the surface and footwear conditions (tile floor with no shoes). Pearson correlations between BKG values from the first and second testing days are shown in Table 1 (column 1). Retest coefficients ranged from .72 to .91, with a mean of .80, indicating good retest reliability.

### Discussion

BKG provides gait cycle data in four conceptually meaningful domains. The findings indicate that there are clear and intuitive consequences for footwear and particularly walking surface on BKG gait metrics, thereby highlighting the importance of standardizing these conditions when assessing gait, as they would otherwise introduce error into the evaluation. BKG data can produce reliable gait data across four conceptual categories over time when the walking surface and footwear are standardized.

Study 2 examines the convergent and predictive validity of the BKG.

## STUDY 2

Evaluates whether BKG gait indices demonstrate convergent validity with another established measure of gait (NIH 4-meter gait), and predictive validity with respect to the endorsement of CDC concussion symptoms. The application to concussion is beneficial, as it represents a growing public health concern [18], and remains the leading global contributor to all disabilities and deaths due to trauma [19]. This study will also examine whether BKG gait compares favorably to other conventional measures of movement (gait and balance) that have previously predicted concussion symptoms [20, 21].

### Methods

#### Patient and Public Involvement

Participants agreed to have their anonymous data aggregated for subsequent analysis to help researchers better understand gait and its relation to important outcomes. Study 2 data were examined archivally, drawing from an extensive and ongoing data collection involving clinical assessments in medical settings following a suspected concussion, and from non-clinical settings where asymptomatic individuals were undergoing baseline testing as a condition of sports participation. The latter draws from community-based leagues and those in collegiate level play (club athletes). Consequently, the sample represents a diverse set of individuals with respect to age and ability.

#### Participants

1,008 individuals (53.4% female) were evaluated across 8 sites, with participants aged 8 to 50 years (M = 16.98, SD = 4.43). Of these individuals, 950 were undergoing standardized preseason baseline assessments, while 58 individuals were undergoing post-concussion evaluations in medical settings. Data were from consecutive evaluations from January 2018 to September 2020, with no exclusions. Participants were not compensated for completing the battery, as it was encouraged by their respective leagues for those completing baselines, and it was part of the medical evaluation for those receiving post concussive care. Gait assessments were standardized and involved walking on a solid surface with no shoes.

#### Procedure

All participants completed the same standardized battery (known as SportGait), delivered on a Microsoft Surface Pro. The SportGait battery is administered in the following order: the CDC concussion symptom checklist, two 20-foot walks while wearing sensors, NIH 4-meter gait test, BESS, and Conner’s Continuous Performance Test (CCPT), 3^rd^ edition. CCPT 3 data were not examined for this study.

The 31-item CDC concussion symptom checklist (each item endorsed as present or absent) includes 11 danger items such as slurred speech, vomiting or nausea, persistent headache, unequal pupil, and 20 less severe symptoms including confusion, sensitivity to light, clumsiness, visual disturbance. Items endorsed were summed to create a single total score, with higher scores denoting a greater number of experienced symptoms.

BKG data from the gait observation were available from all four sensors. To match study 1 data, we here present BKG from the sacrum sensor. Participants and researchers were blind to BKG.

The National Institutes of Health (NIH) 4-Meter Gait test involves taking the fastest of two walks down the 4-m track assessed with an automated timer. The BESS includes six poses on two surfaces, following standardized procedures and the total BESS score was used. All scores were standardized T-scores based on norms for each measure.

### Results

#### Predicting NIH 4-meter gait with the BKG

BKG variables were shown to consistently correlate with NIH 4-m gait scores, with the strongest correlations emerging for stride time and inversely with power (Table 1, column 2). Cumulatively, BKG variables are highly correlated (multiple *R* = .51) with NIH 4-meter gait scores (*R*-square = .256, *F* = 21.4, *df* = 16, 993, *p* < .001). This indicates that BKG variables converge with gait speed.

Regression Analyses to Predict CDC Concussion Symptom Endorsement Using the BKG, NIH 4-meter Gait Test, and BESS

Two hierarchical regressions were performed. For both regressions, age and gender were entered in block 1. In the first regression, NIH 4-M Gait scores were entered in block 2, while for the second regression, BESS total errors were entered in block 2. For both regression analyses, the raw values of the BKG variables established in Study 1 were entered in block 3.

Combined, the covariates of age and gender account for 0.7% variance in CDC symptom endorsement. The NIH 4-M Gait and BESS total errors then add significant incremental variance to the prediction of CDC concussion symptoms, explaining an additional 2.1% variance and 1.7% variance, respectively. Finally, the BKG variables significantly predicted CDC concussion symptoms after accounting for age, gender, NIH 4-m Gait, and BESS scores, explaining an additional 4.9 to 5.7% variance (see Tables 2 and 3). Importantly, BKG variables representing all four gait domains of stride, stability, symmetry, and power were significant predictors.

**Table 2.** Stepwise Linear Regression using NIH 4-M Gait variables to Predict CDC Concussion Symptom Endorsement, Controlling for Age and Gender

| Predictor | *b* | *b* 95% CI<br>[LL,UL] | $\beta$ | *sr* | *r* | Model $R^2$ (Adj $R^2$) | $R^2$<br>Change |
| --- | --- | --- | --- | --- | --- | --- | --- |
| (Intercept) | -1.019 | [-2.68, 0.64] |  |  |  |  |  |
| Age | -0.031** | [-0.05, -0.01] | -0.108 | -0.090 | -0.072 |  |  |
| Gender | -0.038 | [-0.22, 0.15] | -0.015 | -0.034 | -0.047 |  |  |
| <u>Block 1</u> |  |  |  |  |  | 0.007* (0.005) | 0.007* |
| NIH 4M Gait | 0.238** | [0.07, 0.41] | 0.102 | 0.087 | 0.138 |  |  |
| <u>Block 2</u> |  |  |  |  |  | 0.024** (0.021) | 0.017** |
| Stride time | 0.004** | [0.00, 0.01] | 0.129 | 0.081 | 0.093 |  |  |
| Dbl stan | 0.001 | [-0.01, 0.01] | 0.006 | 0.004 | 0.084 |  |  |
| Gait smoothness | -0.352** | [-0.51, -0.20] | -0.136 | -0.127 | -0.130 |  |  |
| Forward power | 0.111 | [-0.68, 0.90] | 0.013 | 0.008 | -0.086 |  |  |
| Side power | 0.655 | [-0.37, 1.68] | 0.064 | 0.039 | -0.071 |  |  |
| Vertical power | 0.078 | [-0.42, 0.58] | 0.017 | 0.009 | -0.073 |  |  |
| Total power | -0.269 | [-1.14, 0.61] | -0.028 | -0.018 | -0.097 |  |  |
| Sway | 10.602* | [0.25, 20.96] | 0.085 | 0.062 | 0.056 |  |  |
| Turnaround sway | -0.673 | [-6.09, 4.74] | -0.009 | -0.007 | 0.055 |  |  |
| Sway velocity | 0.180 | [-1.71, 2.07] | 0.009 | 0.006 | 0.021 |  |  |
| Support var | -0.006 | [-0.02, 0.01] | -0.039 | -0.034 | 0.036 |  |  |
| Striking force | -0.121* | [-0.23, -0.02] | -0.098 | -0.068 | -0.100 |  |  |
| Pushing force | -0.080 | [-0.23, 0.07] | -0.043 | -0.031 | -0.093 |  |  |
| For movement symmetry | -0.005 | [-0.01, 0.00] | -0.048 | -0.041 | -0.012 |  |  |
| Vert movement symmetry | 0.008* | [0.00, 0.01] | 0.089 | 0.078 | 0.039 |  |  |
| Vert sway symmetry | -0.007 | [-0.02, 0.00] | -0.061 | -0.043 | -0.006 |  |  |
| <u>Block 3</u> |  |  |  |  |  | 0.073** (0.055) | 0.049** |

*Note.* A significant *b*-weight indicates that the beta weight and semi-partial correlation are also significant. *b* represents unstandardized regression weights, $\beta$ indicates the standardized regression weights, $sr^2$ represents the semi-partial correlation squared, *r* represents the zero-order correlation, *LL* and *UL* indicated the lower and upper limits of a confidence interval, respectively. BKG variables represent the four gait domains of stride, stability, symmetry, and power. When the BKG variables were examined independently (without entering NIH 4 m gait and BESS scores first), the BKG variables could account for over 5% of the variance (Adjusted r-square = .053, $F = 8.032$ , $p < .01$ ). The variables stepping into this regression equation were Stride time, Gait smoothness, Striking force, Vertical sway symmetry, sway, and Vertical movement symmetry. Age was also a significant predictor. \* Indicates $p < .05$ , \*\* indicates $p < .01$ .

**Table 3.** Stepwise Linear Regression using BESS Total Errors and BKG variables to Predict CDC Concussion Symptom Endorsement, Controlling for Age and Gender

| Predictor | *b* | *b* 95% CI<br>[LL,UL] | $\beta$ | *sr* | *r* | Model $R^2$ (Adj<br>$R^2$) | $R^2$<br>Change |
| --- | --- | --- | --- | --- | --- | --- | --- |
| (Intercept) | -0.635 | [-2.21, 0.94] |  |  |  |  |  |
| Age | -0.036** | [-0.06, -0.01] | -0.126 | -0.105 | -0.072 |  |  |
| Gender | -0.020 | [-0.38, -0.16] | -0.008 | -0.007 | -0.047 |  |  |
| <u>Block 1</u> |  |  |  |  |  | 0.007* (0.005) | 0.007* |
| BESS Total | 0.028** | [0.01, 0.04] | 0.113 | 0.108 | 0.120 |  |  |
| <u>Block 2</u> |  |  |  |  |  | 0.020** (0.017) | 0.013** |
| Stride time | 0.004** | [0.00, 0.01] | 0.141 | 0.088 | 0.093 |  |  |
| Double stance | 0.002 | [-0.01, 0.01] | 0.018 | 0.013 | 0.084 |  |  |
| Gait smoothness | -0.323** | [-0.48, -0.17] | -0.145 | -0.124 | -0.130 |  |  |
| Forward power | 0.167 | [-0.62, 0.96] | 0.013 | 0.011 | -0.086 |  |  |
| Side power | 0.404 | [-0.61, 1.42] | 0.040 | 0.024 | -0.071 |  |  |
| Vertical power | -0.064 | [-0.56, 0.43] | -0.014 | -0.008 | -0.073 |  |  |
| Total power | -0.381 | [-1.25, 0.49] | -0.040 | -0.026 | -0.097 |  |  |
| Sway | 11.303* | [0.99, 21.62] | 0.091 | 0.066 | 0.056 |  |  |
| Turnaround sway | -1.288 | [-6.69, 4.12] | -0.018 | -0.014 | 0.055 |  |  |
| Sway velocity | -0.028 | [-1.92, 1.86] | -0.001 | -0.001 | 0.021 |  |  |
| Support variability | -0.007 | [-0.02, 0.01] | -0.044 | -0.038 | 0.036 |  |  |
| Striking force | -0.117* | [-0.22, -0.01] | -0.095 | -0.066 | -0.100 |  |  |
| Pushing force | -0.070 | [-0.22, 0.08] | -0.038 | -0.028 | -0.093 |  |  |
| Forward movement symmetry | -0.007 | [-0.02, 0.00] | -0.059 | -0.050 | -0.012 |  |  |
| Vertical movement symmetry | 0.008** | [0.00, 0.01] | 0.091 | 0.080 | 0.039 |  |  |
| Vertical sway symmetry | -0.006 | [-0.02, 0.00] | -0.054 | -0.038 | -0.006 |  |  |
| <u>Block 3</u> |  |  |  |  |  | 0.077** (0.059) | 0.057** |

*Note.* A significant *b*-weight indicates that the beta weight and semi-partial correlation are also significant. *b* represents unstandardized regression weights, $\beta$ indicates the standardized regression weights, $sr^2$ represents the semi-partial correlation squared, *r* represents the zero-order correlation, *LL* and *UL* indicated the lower and upper limits of a confidence interval, respectively. BKG variables represent the three gait domains of stride, stability, and power, respectively. When both the BESS and NIH 4M Gait scores were entered together in block 2, they predicted 3.3% of the variance in total symptoms (Adjusted r-square = .033, $F=14.92$ , $p < .01$ ). BKG variables added in block 3 then added 4.7% of variance to the model predicting total symptoms (Adjusted r-square = .065, $F=6.87$ , $p < .01$ ). \* indicates $p < .05$ , \*\* indicates $p < .01$ .

## General Discussion

Tao and colleagues [4] reviewed gait measurements in clinical settings using a variety of technologies (including wearable accelerometers), illustrating considerable diversity in how to quantify gait. Accelerometers can identify and derive specific gait cycle features [22, 23], that correspond to mTBIs [24].

The current studies add to this research by not only showing that reliable gait values can be extracted from the BKG for sixteen variables representing four meaningful categories (stride, stability, symmetry, and power), but that BKG scores are also sensitive to extraneous factors such as footwear and especially walking surface. Thus, the standardization of gait assessments with respect to footwear and walking surface is critical to avoid confounding variables when interpreting gait. This is especially relevant in clinical settings, where the interpretation of gait data has potential screening, diagnostic, monitoring, and therapeutic implications, and where strict control over footwear and the walking surface can be achieved.

BKG variables also predict gait speed, indicating convergence with one of the more longstanding measures of gait that has been linked to important outcomes. BKG variables were also shown to out-predict the separate and combined effects of two frequently used and well-validated measures of gait and balance (NIH 4-m gait and BESS scores), and BKG also adds significantly to the prediction of CDC concussion symptoms over these measures. Given the brevity of the BKG assessment, this is an especially important finding; suggesting that although gait speed is a good predictor, sensor-based assessments of motion provide even more useful data, which is in keeping with the literature on mTBI [24].

The goal of validating the BKG gait assessment is to identify unique motion signatures to establish objective biomarkers of health and illness. The long-term objective is to identify diagnostic biokinetic signatures for various vascular, neurological, and orthopedic conditions, and use BKG to monitor therapeutic response or disease progression. BKG measurement can also be part of a baseline screening protocol to facilitate the detection of exercise-related injuries and for determining when it is safe to return to activity.

This biokinetic knowledge could contribute to innovative measures of health, identification of pre-disease pathways, and novel monitoring strategies. Other studies have reported associations between accelerometer-assessed gait for geriatric syndromes such as falls and frailty [25, 26] and neurologic conditions such as Alzheimer’s Disease, Parkinson’s Disease, and stroke [27, 28, 29]. We add to the literature by demonstrating the prediction of concussion symptoms over and above what is predicted by gait speed and balance. BKG data may be especially useful when patients are either unable (e.g., children or adolescents, [40]) or unwilling (e.g., athletes wanting to return prematurely, [41]) to accurately report concussion symptoms.

BKG data for studies 1 and 2 were derived from a single sacrum sensor. Although a richer dataset could be examined from sensors located at the wrist, ankles, and sacrum, our rationale for adopting this simplified analysis is to mimic single sensor approaches using the technology available in smartphones [30], which could result in broader adoption of the BKG.

### Conclusions

The current research documents that an analytic gait process (BKG) shows robust test-retest reliability and sensitivity to extraneous factors of footwear and surface type (Study 1). Moreover, BKG variables can predict gait speed, and self-reported concussion symptoms to a greater degree than previously validated measures of gait and balance (Study 2).

### Clinical Implications

Two studies provide important initial steps in validating the BKG and its applicability to concussion, illustrating that a brief automated analysis of gait can achieve reliability and outperform established measures with respect to a concussion-specific outcome. Future research will explore additional BKG variables, establish normative data for various health conditions, and explore data collection using a mobile platform.

### Limitations

Limitations include not having clinical diagnoses to validate the experience of concussion, the absence of data from older adults, and lacking information regarding comorbid health conditions.

## Data Availability

All data produced in the present study are available upon reasonable request to the first author

