## Supplemental Table 1 for "Reliability and Predictive Validity of a Gait Assessment using Inertial Measurement Units: The Importance of Standardizing Walking Surface and Footwear"

**Supplemental Materials**

**Table 1**

*Fixed Effects ANOVA assessing the effects of Footwear and Surface on BKG Variables*

| Dependent Variable | *Factor* | *Sum of Squares* | *df* | *Mean Square* | *F* | *d* |
|---|---|---|---|---|---|---|
| Total power | Intercept | 55.04 | 1 | 55.04 | 4231.18** | 8.63 |
| | Surface | 0.69 | 1 | 0.69 | 52.71** | 0.97 |
| | Footwear | 0.01 | 1 | 0.01 | 0.65 | 0.11 |
| | Surface * Footwear | 0.02 | 1 | 0.02 | 1.69 | 0.17 |
| | Error | 2.94 | 226 | 0.01 | | |
| | Corrected Total | 58.52 | 230 | | | |
| Side power | Intercept | 43.52 | 1 | 43.52 | 1825.29** | 5.69 |
| | Surface | 0.35 | 1 | 0.35 | 14.45** | 0.51 |
| | Footwear | 0.00 | 1 | 0.00 | 0.04 | 0.00 |
| | Surface * Footwear | 0.01 | 1 | 0.01 | 0.33 | 0.063 |
| | Error | 5.39 | 226 | 0.02 | | |
| | Corrected Total | 49.16 | 230 | | | |
| Vertical power | Intercept | 262.21 | 1 | 262.21 | 4349.42** | 8.81 |
| | Surface | 3.86 | 1 | 3.86 | 63.96** | 1.07 |
| | Footwear | 0.60 | 1 | 0.60 | 9.99** | 0.42 |
| | Surface * Footwear | 0.10 | 1 | 0.10 | 1.60 | 0.17 |
| | Error | 13.63 | 226 | 0.06 | | |
| | Corrected Total | 279.26 | 230 | | | |
| Forward power | Intercept | 107.68 | 1 | 107.68 | 6425.44** | 10.66 |
| | Surface | 0.46 | 1 | 0.46 | 27.33** | 0.70 |
| | Footwear | 0.01 | 1 | 0.01 | 0.40 | 0.09 |
| | Surface * Footwear | 0.00 | 1 | 0.00 | 0.05 | 0.00 |
| | Error | 3.79 | 226 | 0.02 | | |
| | Corrected Total | 111.72 | 230 | | | |
| Stride time | Intercept | 64945432.86 | 1 | 64945432.86 | 60703.90** | 31.56 |
| | Surface | 6187.35 | 1 | 6187.35 | 5.78* | 0.32 |
| | Footwear | 8604.84 | 1 | 8604.84 | 8.04** | 0.38 |
| | Surface * Footwear | 1100.56 | 1 | 1100.56 | 1.03 | 0.14 |
| | Error | 241791.19 | 226 | 1069.87 | | |
| | Corrected Total | 65210266.20 | 230 | | | |
| Gait smoothness | Intercept | 1294.61 | 1 | 1294.61 | 4616.62** | 9.01 |
| | Surface | 5.64 | 1 | 5.64 | 20.12** | 0.60 |
| | Footwear | 0.10 | 1 | 0.10 | 0.35 | 0.09 |
| | Surface * Footwear | 0.06 | 1 | 0.06 | 0.23 | 0.06 |

| | | | | | | |
|---|---|---|---|---|---|---|
| | Error | 63.38 | 226 | 0.28 | | |
| | Corrected Total | 1367.40 | 230 | | | |
| Turnaround sway | Intercept | 0.79 | 1 | 0.79 | 2573.27** | 6.74 |
| | Surface | 0.00 | 1 | 0.00 | 8.80** | 0.39 |
| | Footwear | 0.00 | 1 | 0.00 | 3.19 | 0.24 |
| | Surface * Footwear | 0.00 | 1 | 0.00 | 0.00 | 0.00 |
| | Error | 0.07 | 226 | 0.00 | | |
| | Corrected Total | 0.86 | 230 | | | |
| Double stance | | | | | | |
| | Intercept | 1577679.78 | 1 | 1577679.80 | 12672.67** | 14.77 |
| | Surface | 1.55 | 1 | 1.55 | 0.01 | 0.00 |
| | Footwear | 810.12 | 1 | 810.12 | 6.50* | 0.34 |
| | Surface*Footwear | 143.24 | 1 | 143.24 | 1.15 | 0.14 |
| | Error | 28135.80 | 226 | 124.50 | | |
| | Corrected Total | 1607586.05 | 230 | | | |
| Striking force | | | | | | |
| | Intercept | 4015.79 | 1 | 4015.79 | 4346.11** | 8.81 |
| | Surface | 5.22 | 1 | 5.22 | 5.65** | 0.31 |
| | Footwear | 6.16 | 1 | 6.16 | 6.67** | 0.35 |
| | Surface * Footwear | 0.18 | 1 | 0.18 | 0.20 | 0.06 |
| | Error | 208.82 | 226 | 0.92 | | |
| | Corrected Total | 4232.05 | 230 | | | |
| Pushing force | | | | | | |
| | Intercept | 2264.56 | 1 | 2264.56 | 6083.72** | 10.35 |
| | Surface | 17.91 | 1 | 17.91 | 48.12** | 0.92 |
| | Footwear | 0.15 | 1 | 0.15 | 0.39 | 0.09 |
| | Surface * Footwear | 0.18 | 1 | 0.18 | 0.49 | 0.09 |
| | Error | 84.12 | 226 | 0.37 | | |
| | Corrected Total | 2361.15 | 230 | | | |
| Forward movement symmetry | | | | | | |
| | Intercept | 849414.75 | 1 | 849414.75 | 8679.79** | 12.49 |
| | Surface | 4983.35 | 1 | 4983.35 | 50.92** | 0.95 |
| | Footwear | 117.04 | 1 | 117.04 | 1.20 | 0.14 |
| | Surface * Footwear | 99.31 | 1 | 99.31 | 1.02 | 0.13 |
| | Error | 22116.63 | 226 | 97.86 | | |
| | Corrected Total | 874959.88 | 230 | | | |
| Vertical sway symmetry | Intercept | 50899.38 | 1 | 50899.38 | 811.12 | 3.79 |
| | Surface | 157.45 | 1 | 157.45 | 2.51 | 0.21 |
| | Footwear | 256.81 | 1 | 256.81 | 4.09* | 0.27 |
| | Surface * Footwear | 65.33 | 1 | 65.33 | 1.04 | 0.14 |

| | | | | | | |
|---|---|---|---|---|---|---|
| | Error | 14181.93 | 226 | 62.75 | | |
| | Corrected Total | 65702.55 | 230 | | | |
| Sway velocity | Intercept | 17.62 | 1 | 17.62 | 3043.67** | 7.35 |
| | Surface | 0.09 | 1 | 0.09 | 14.78** | 0.51 |
| | Footwear | 0.00 | 1 | 0.00 | 0.05 | 0.00 |
| | Surface * Footwear | 0.00 | 1 | 0.00 | 0.05 | 0.00 |
| | Error | 1.31 | 226 | 0.01 | | |
| | Corrected Total | 18.98 | 230 | | | |

*Note.* To conserve space, only analyses resulting in significant effects are displayed here. * indicates $p < .05$, ** indicates $p < .01$. $d$ indicates Cohen's $d$ as a measure of effect size. Thirteen of the sixteen BKG outputs resulted in significant effects (11 for surface, 5 for footwear, with three having both). When combined with the effect sizes, the concerns for alpha inflation are minimal.
